# Meta-topologies define distinct anatomical classes of brain tumors linked to histology and survival

**DOI:** 10.1101/2021.11.20.21266624

**Authors:** Julius M. Kernbach, Daniel Delev, Georg Neuloh, Hans Clusmann, Danilo Bzdok, Simon B. Eickhoff, Victor E. Staartjes, Flavio Vasella, Michael Weller, Luca Regli, Carlo Serra, Niklaus Krayenbühl, Kevin Akeret

**Author notes:** Corresponding author:* Kevin Akeret, MD, Department of Neurosurgery, Clinical Neuroscience Center, University Hospital and University of Zurich, Frauenklinikstrasse 10, CH-8091 Zurich, Switzerland. shared (Julius M. Kernbach, Daniel Delev).

## Abstract

**Background:** The current WHO classification integrates histological and molecular features of brain tumors. The aim of this study was to identify generalizable topological patterns with the potential to add an anatomical dimension to the classification of brain tumors.

**Methods:** We applied non-negative matrix factorization as an unsupervised pattern discovery strategy to the fine-grained topographic tumor profiles of 936 patients with primary and secondary brain tumors. From the anatomical features alone, this machine learning algorithm enabled the extraction of latent topological tumor patterns, termed *meta-topologies*. The optimal parts-based representation was automatically determined in 10,000 split-half iterations. We further characterized each meta-topology’s unique histopathologic profile and survival probability, thus linking important biological and clinical information to the underlying anatomical patterns

**Results:** In primary brain tumors, six meta-topologies were extracted, each detailing a transpallial pattern with distinct parenchymal and ventricular compositions. We identified one infratentorial, one allopallial, three neopallial (parieto-occipital, frontal, temporal) and one unisegmental meta-topology. Each meta-topology mapped to distinct histopathologic and molecular profiles. The unisegmental meta-topology showed the strongest anatomical-clinical link demonstrating a survival advantage in histologically identical tumors. Brain metastases separated to an infra- and supratentorial meta-topology with anatomical patterns highlighting their affinity to the cortico-subcortical boundary of arterial watershed areas.

**Conclusions:** Using a novel data-driven approach, we identified generalizable topological patterns in both primary and secondary brain tumors Differences in the histopathologic profiles and prognosis of these anatomical tumor classes provide insights into the heterogeneity of tumor biology and might add to personalized clinical decision making.

## Introduction

Each year, over 300,000 people worldwide are diagnosed with brain tumors, which cause more than 200,000 deaths and 7,600,000 disability-adjusted life years.^1^ In recent decades, major advances in the histologic and molecular profiling of brain tumors have been achieved and implemented into classification systems and diagnostic guidelines.^2,3^ The most recent WHO Classification of Tumors of the Central Nervous System further strengthens this integration of molecular and histological parameters.^4,5^ The anatomical phenotype of brain tumors, however, is of minor relevance in this classification. Previous reports indicate an association between tumor location and biological tumor signature.^6–9^ We propose a unified data-driven framework tailored to identify generalizable topological patterns, which may enhance our understanding and the classification of brain tumors.^8,9^

Specific anatomical patterns are found in numerous neurological diseases. Neurodegenerative disorders differ in their atrophy patterns^10^, or autoinflammatory diseases preferentially affect distinct CNS structures.^11^ The molecular mechanisms behind this selective vulnerability of the brain, referred to as pathoclisis,^12^ remain largely elusive. In brain tumors, descriptions of topographic prevalence and relative spatial density also implicate differences in the anatomical phenotype.^8,9^ Defining anatomical classes of brain tumors requires the identification of topological relationships that are consistent across patients. Topographic analyses can provide a description of the involvement of individual anatomical structures. However, analyzing each neuroanatomical location as a single unit deters intuitive interpretation by omitting interaction effects between spatially adjacent or distant areas.

In contrast, studies on topology aim to incorporate the relative positions of the individual anatomical components to each other.^13^ We propose a novel framework tailored to identify generalizable patterns in brain tumor topology using non-negative matrix factorization (NNMF).^14^ Factorization methods, including NNMF, are often applied in the analyses of genomic signatures across various cancer types^15– 17^ and provide unique advantages for the purpose of the present study. The inherent non-negative constraint of the applied algorithm enables an intuitive and direct interpretation of the derived patterns. In contrast, alternative dimensionality reduction tools such as principal component analysis would hurt intuitive interpretation of any distributed effects by recovering patterns through incomprehensible combinations of positive and negative cancellations of the extracted low-dimensional patterns.^14^ Further, NNMF is considered a sum-of-parts approach. It purposefully can appreciate the mutual functional dependence between individual neuroanatomical locations and enables an interpretation on a topological level.

In the present computational study, we design an unsupervised data-led approach using machine learning to identify an optimal factorization of latent neuroanatomical meta-patterns in brain tumors that we henceforth call *meta-topologies*. First, we extract a low-dimensional embedding from the fine-grained neuroanatomical distributions using unsupervised pattern discovery. Second, we assess the subject-specific expression of the derived tumor configurations for their histopathologic identity and prognostic relevance. By introducing brain tumor meta-topologies, we intend to supplement our biological understanding and the individual profiling of brain tumors to inform individualized treatment decisions and assist in tailored therapy customized to the single patient.^18,19^

## Materials and Methods

This study was approved by the ethical review board of the Canton of Zurich, Switzerland (KEK ZH 01120). Reporting of results is in accordance with the STROBE statement.^20^

### Data source

Topographic tumor profiles were obtained from a previously published and openly available single-center cohort of *n* = 1000 consecutive patients with newly diagnosed primary or secondary brain tumors (https://doi.org/10.5281/zenodo.5457402).^9^ The eligibility criteria comprised: (i) first diagnosis with consecutive histopathologic confirmation of a primary or secondary brain tumor; (ii) no pretreatment or previous cranial surgery; (iii) intraparenchymal encephalic tumor location; (iv) availability of preoperative MRI data. We excluded 44 patients with primary central nervous system lymphoma due to considerations of statistical power and 20 tumors with indistinct gyral patterns for the analyses in this study. Each patient’s topographic tumor profile was based on a standardized whole-brain parcellation protocol^21^ and contained 120 anatomical annotations (Supplemental Figure 2).

### Data-led deconvolution of hidden tumor meta-topologies

We sought to explore coherent topological anatomical patterns that may be hidden in the rich spatial descriptions of the topographic phenotypes. We capitalized on NNMF as a lossy multivariate pattern discovery strategy.^14^ This unsupervised machine learning algorithm can identify the form and patient-specific combination of latent topological patterns that together compose the individual neuroanatomical tumor phenotypes. These derived sum-of-parts representations are henceforth called *meta-topologies*. More formally, NNMF achieves a low-rank approximation of the data *V*, with *V* reflecting the 120 topographic summaries, with dimensions of *m* x *n* (*m* = number of anatomical items, *n* = number of patients), by partitioning the interindividual variation in anatomical items into a basic matrix *W* of *k* parts-based factor representations. The matrix of the latent factor loadings *H* indicated how relevant each emerging meta-topology was to describe an individual patient’s tumor phenotype. Accordingly, *W* and *H* carried *m* x *k* and *k* x *n* dimensions, respectively. Given by *V = WH*, the latent factorization decomposed the actual tumor phenotype in a particular patient into a part-based representation.

### Optimal factorization based on quantitative model evaluation

To find the optimal parts-based representation, we capitalized on robustness measures and the ability to generalize to new populations. We applied a data-driven out-of-sample evaluation strategy to determine the most robust and generalizable representations of tumor meta-topologies across 10,000 bootstrapped split-half iterations. We quantitatively assessed the resulting factorizations for (a) generalizability by measuring the increase in out-of-sample reconstruction error (RE)^22^ and (b) stability using the adjusted Rand Index (aRI).^23,24^ The optimal factorization should be reflected in both a lower increase in out-of-sample RE and a higher aRI in the majority of the iterations.

We quantified the out-of-sample RE by projecting the data of one half onto the latent dimensions from the other half. The RE represents the absolute difference between the reconstructed and original matrix. Accordingly, the increase in out-of-sample RE demonstrates how much worse the data matrix is reconstructed by the basis matrix obtained from the model-unseen sample compared to the basis matrix recovered from the within-sample split. A lower increase in out-of-sample RE compared with the within-sample RE hence indicates better generalizability.

Stability was assessed using the aRI. As a modified version of the Rand Index, the aRI is stricter and allows for improved discrimination.^23,24^ The aRI is adjusted for chance; that is, the aRI penalizes for the placement of two data points from different true clusters into the same cluster. The aRI is ensured to have values close to 0 for random labeling and yields a value between -1 and +1, with negative values when the index is less than the expected index. In this study, the aRI was used to measure the correspondence between the factorizations derived from the two split-samples based on the assignment of the anatomical items to the parts-based meta-topologies. Higher values of aRI indicate better correspondence between the two factorizations derived in separate split-half realizations, and a value of 1 represents an identical assignment.

We selected the most generalizable and robust factorization with rank *k* when the mean increase in out-of-sample RE of the all 10,000 split-half realizations was minimized, while the mean aRI was maximized.

### Statistical analysis

The Kaplan Meier method was used to estimate survival probabilities of tumor meta-topologies, and the log-rank test with Bonferroni-Holm correction for multiple testing was applied for pairwise comparisons.^25^ Multivariable survival analysis was performed using the Cox proportional hazards regression model.^26,27^ Unadjusted models were compared to models with adjustment for histopathologic tumor subtype. Given the exploratory nature of this study, the results were interpreted based on the level of evidence without the definition of a level of statistical significance: p < 0.001: very strong evidence; p < 0.01: strong evidence; p < 0.05 evidence; p < 0.1 weak evidence; p > 0.1: no evidence.^28^

### Code availability

Data analyses were conducted in Python 3.8.5 (IPython 7.21.0) and R 4.0.0 (RStudio 1.3.1093) environments. Full codes are available online (https://doi.org/10.5281/zenodo.5515356).

## Results

Detailed demographic, histopathologic, and clinical cohort characteristics are provided in Supplemental Table 1. We quantitatively assessed the most robust and generalizable NNMF factorization for primary brain tumors and brain metastases separately by combining the maximized mean aRI and the out-of-sample RE with the least relative increase (Supplemental Figure 1). We extracted the optimal factorization as *k* = 6 meta-topologies in primary brain tumors (Supplemental Figure 1A) and *k* = 2 in brain metastases (Supplemental Figure 1B).

### Meta-topologies in primary brain tumors

We identified six meta-topologies in primary brain tumors. Each meta-topology showed a distinct composition of anatomical items (Figure 1, Supplemental Figure 2A, Supplemental Table 2) and level of gyrality (Supplemental Table 3) that informed post-hoc naming. Meta-topologies 1-5 predominantly reflected supratentorial anatomy. Infratentorial structures were captured in meta-topology 6. Distinct combinations of gyral, ventricular, and radial tumor anatomy characterized the unique constellations of meta-topologies 1-5: we identified three meta-topologies with neopallial mapping (parieto-occipital, frontal, temporal), one with predominantly allopallial enrichment and one meta-topology lacking a gyral pattern (unisegmental).

**Figure 1.**
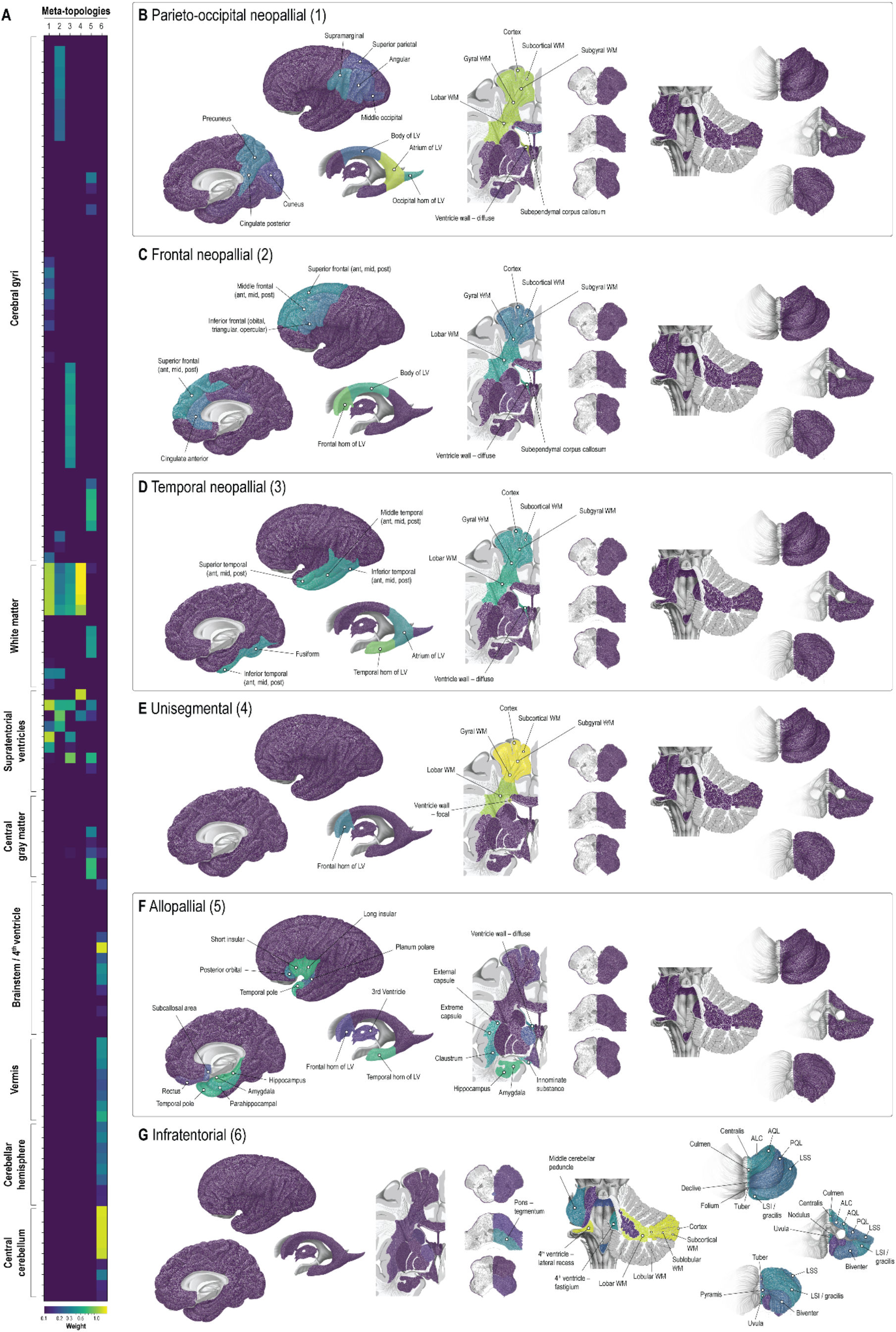
Meta-topologies in primary brain tumors. **A**. Meta-topologies in primary brain tumors with their respective anatomical configurations. A detailed description of the anatomical distributions is provided in Supplemental Figure 2. **B-G**. Spatial visualizations of three neopallial (1: parieto-occipital; 2: frontal; 3: temporal), unisegmental (4), allopallial (5), and infratentorial (6) meta-topologies. The anatomical items with the highest differential weight are labeled. *Abbreviations:* ALC, ala lobuli centralis; Ant, anterior third; AQL, anterior quadrangular lobule; LSI, inferior semilunar lobule; LSS, superior semilunar lobule; LV, lateral ventricle; mid, middle third; post, posterior third; PQL, posterior quadrangular lobule; WM, white matter.

#### *Parieto-occipital neopallial primary brain tumors* (meta-topology 1, Figure 1B)

Most relevant anatomical items included the atrium of the lateral ventricle (loading weight 1.19), diffuse involvement of the wall of the lateral ventricle (1.17), all cerebral white matter sectors (lobar and gyral 1.11, subgyral 1.10, subcortical 1.09) and the cerebral cortex (1.07). In addition, meta-topology 1 mapped to the occipital horn and the body of the lateral ventricle, the subependymal corpus callosum, and gyri of the parieto-occipital neopallium (supramarginal, precuneus, angular, middle occipital, superior parietal, cuneus). Primary brain tumors characterized primarily by the parieto-occipital neopallial meta-topology 1 (*n* = 141) were multigyral in 46.1% (*n* = 65), unigyral in 49.6% (*n* = 70), and affected no cerebral gyrus in 4.3% (*n* = 6)

#### *Frontal neopallial primary brain tumors* (meta-topology 2, Figure 1C)

The frontal horn of the lateral ventricle featured the highest loading weight (0.82), followed by the body of the lateral ventricle (0.52), diffuse involvement of the wall of the lateral ventricle (0.52), and the cerebral lobar white matter sector (0.43). Meta-topology 2 was located in the gyral, subgyral, and subcortical white matter sectors, the cerebral cortex, and gyri of the fronto-medial neopallium (superior, middle, and inferior frontal gyri, anterior cingulate gyrus). Primary brain tumor characterized by the frontal neopallial meta-topology 2 (*n* = 115), showed a multigyral involvement in 57.4% (*n* = 66), unigyral in 27.8% (*n* = 32), and no gyral involvement in 14.8% (*n* = 17).

#### *Temporal neopallial primary brain tumors* (meta-topology 3, Figure 1D)

Meta-topology 3 was uniquely defined by the temporal horn of the lateral ventricle (0.86), diffuse involvement of the wall of the lateral ventricle (0.53), and the cerebral lobar white matter sector (0.52). Meta-topology 3 also mapped to the other cerebral radial sectors (gyral, subgyral, subcortical), the cerebral cortex, and temporal neopallial gyri (superior, middle, and inferior temporal, fusiform). The atrium of the lateral ventricle was also affected, yet to a lesser extent than the temporal horn. Primary brain tumors mapping predominantly on the temporal neopallial meta-topology 3 (*n* = 69) were multigyral in 56.5% (*n* = 39), unigyral in 39.1% (*n* = 27) and involved no gyrus in 4.3% (*n* = 3).

#### *Unisegmental primary brain tumors* (meta-topology 4, Figure 1E)

Meta-topology 4 represented supratentorial primary brain tumors mapping to the cerebral cortex (1.58) and all cerebral white matter sectors (subcortical 1.57, subgyral 1.54, gyral 1.41, lobar 1.07). Ventricular involvement was primarily limited to focal contact to the wall of the lateral ventricle (1.25), specifically in the frontal horn of the lateral ventricle (0.30). Gyral constellations showed no relevant contribution to meta-topology 4. Notably, primary brain tumors belonging to meta-topology 4 (*n* = 179) were dominated by a pronounced unigyral character (82.1%, *n* = 147; 17.9 % (*n* = 32) multigyral).

#### *Allopallial primary brain tumors* (meta-topology 5, Figure 1F)

Meta-topology 5 was dominated by the amygdala (0.65), the hippocampus (0.62), long (0.61) and short (0.60) insular gyri, the temporal horn of the lateral ventricle (0.57), temporal pole (0.56), parahippocampal gyrus (0.47), innominate substance (0.44), as well as extreme (0.43) and external (0.41) capsules. Meta-topology 5 additionally mapped to the claustrum, posterior orbital gyrus, subcallosal area, planum polare, and thalamus, as well as the frontal horn of the lateral ventricle and the third ventricle. Diffuse involvement of the wall of the lateral ventricle added more relevantly to the constellation of the meta-topology than a focal ventricular contact. Primary brain tumors mapping predominantly on the allopallial meta-topology 5 (*n* = 70) were multigyral in 55.7% (*n* = 39), unigyral in 12.9% (*n* = 9) and involved no gyrus in 31.4% (*n* = 22).

#### *Infratentorial primary brain tumors* (meta-topology 6, Figure 1G)

The last meta-topology was determined by infratentorial structures: the cerebellar cortex (1.37), the lateral recess of the fourth ventricle (1.35), as well as the cerebellar white matter sectors (subcortical 1.34, sublobular 1.34, lobular 1.33, lobar 1.27). Both the vermian and the hemispheric lobules contributed to the neuroanatomical constellation of meta-topology 6. Of the primary brain tumors matching best on meta-topology 6 (*n* = 72), 98% (*n* = 71) did not involve a cerebral gyrus. There was only one tumor (1.4%) with multigyral anatomy.

Collectively, all identified meta-topologies in primary brain tumors involved distinct ventricular segments and shared a characteristic transpallial pattern. Each supratentorial meta-topology presented a distinct gyral pattern except for meta-topology 4, which was predominantly unigyral in nature. The unigyral character of meta-topology 4 was uniquely associated with a strong contribution of a focal contact to the ventricle wall, while all other supratentorial meta-topologies depicted a diffuse ventricular involvement.

### Meta-topologies in brain metastases

We quantitatively identified two meta-topologies in brain metastases (Figure 2, Supplemental Figure 2B, Supplemental Table 4). A separation between infratentorial (meta-topology 1) and supratentorial (meta-topology 2) topological patterns emerged.

**Figure 2.**
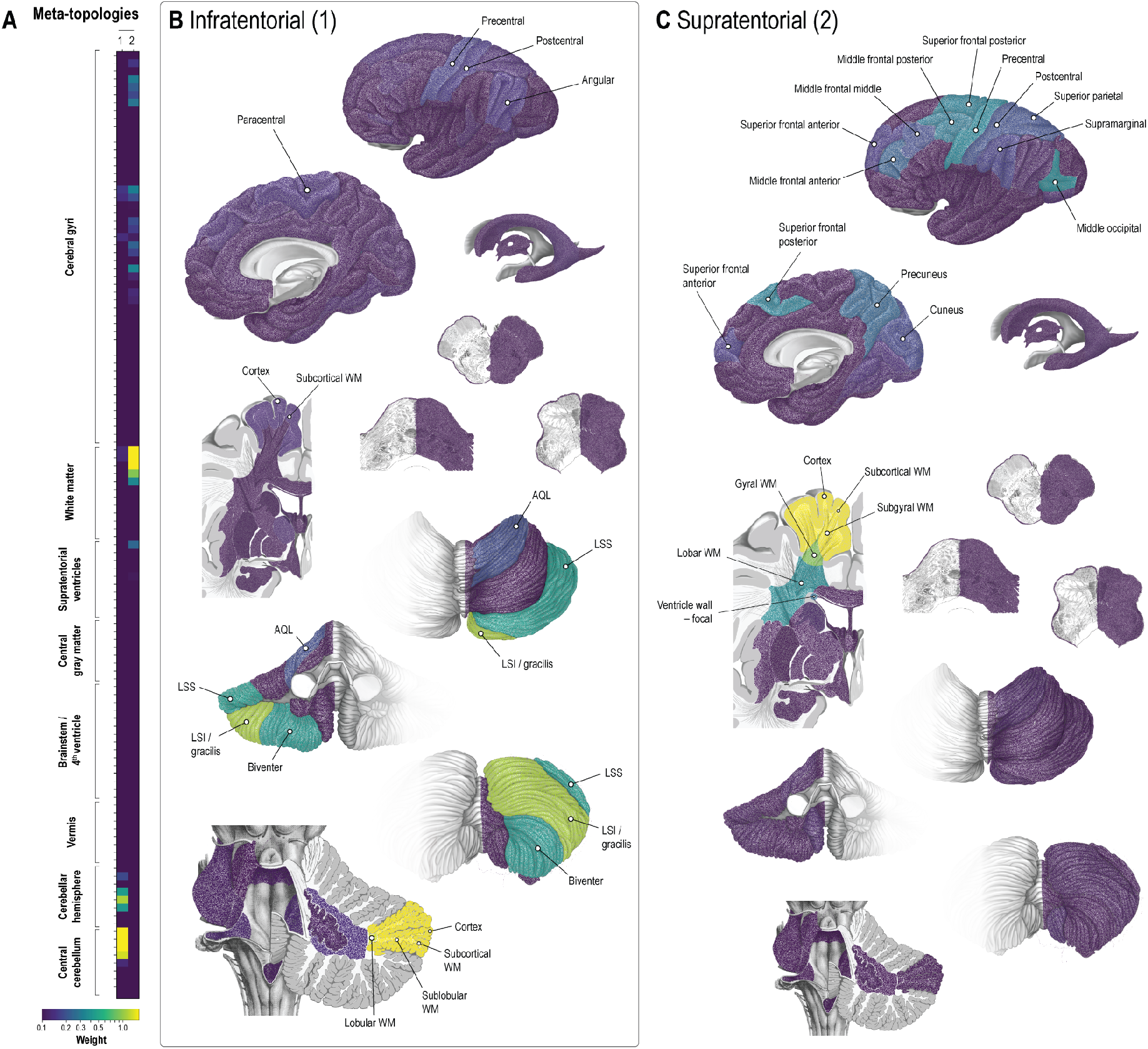
Meta-topologies in brain metastases. **A**. Meta-topologies in brain metastases with their respective anatomical configurations. A detailed description of the anatomical distributions is provided in Supplemental Figure 2. **B-C**. Spatial visualization of the infra-(1) and supratentorial (2) meta-topologies. The anatomical items with the highest differential weight are labeled. *Abbreviations:* AQL, anterior quadrangular lobule; LSI, inferior semilunar lobule; LSS, superior semilunar lobule; LV, lateral ventricle; PQL, posterior quadrangular lobule; WM, white matter.

#### *Infratentorial brain metastases* (meta-topology 1, Figure 2B)

The topological pattern was dominated by the cerebellar cortex (2.18) and the superficial cerebellar white matter sectors (subcortical 2.17, sublobular 1.94, lobular 1.49). In addition, meta-topology 1 mapped strongly to the inferior semilunar/gracilis lobule (1.24) and, although less pronounced, to the superior semilunar (0.47), biventer (0.45), and anterior quadrangular (0.19) lobules. There was also discrete enrichment in specific supratentorial items, such as the precentral, postcentral, paracentral, angular gyri, and the cerebral cortex and cerebral subcortical white matter sector.

#### *Supratentorial brain metastases* (meta-topology 2, Figure 2C)

Meta-topology 2 was again determined by superficial structures, namely the cerebral cortex (2.83) and cerebral subcortical white matter sector (2.83), followed by the cerebral subgyral white matter sector (2.30). Weaker enrichment was seen in the cerebral gyral (1.03) and lobar (0.37) white matter sectors. On a gyral level, we found the strongest factor contribution in the middle occipital (0.36), precentral (0.34), posterior third of the superior frontal (0.32), and posterior third of the middle frontal (0.30) gyri. Meta-topology 2 also mapped to the anterior third of the superior frontal, anterior two-thirds of the middle frontal, the postcentral and supramarginal gyri, the superior parietal lobule, precuneus, and cuneus. We observed focal contact to the wall of the lateral ventricle (0.26) but no diffuse involvement.

In summary, both meta-topologies in brain metastases showed a strong affinity to the cortex (cerebellar or cerebral) and superficial white matter sectors (especially subcortical). The observed cerebellar lobular and cerebral gyral patterns were consistent with the expected anatomical locations of arterial border zones, i.e., watershed areas. The gyral pattern seen in meta-topology 2 was weakly represented in meta-topology 1, likely due to synchronous supra- and infratentorial metastases. Unlike in primary brain tumors, ventricular segments and deep white matter sectors were not associated with meta-topologies in brain metastases.

### Brain tumor meta-topologies map to distinct histopathologic and molecular profiles

The dominant histopathologic entities in primary brain tumors were glioblastoma (60.7%) and gliomas of WHO grade 3 (16.8%) and 2 (8.0%) (Figure 3A). The histopathologic entities spread differently to the identified meta-topologies in primary brain tumors. Glioblastoma, WHO grade 3 and 2 gliomas, and developmental tumors mapped primarily to supratentorial meta-topologies. Ependymoma appeared in both supra- and infratentorial meta-topologies, while pilocytic astrocytoma and medulloblastoma were almost exclusively associated with the infratentorial pattern (meta-topology 6). Glioblastoma mapped to all neopallial (parieto-occipital (1), frontal (2), and temporal (3)), and the unisegmental (4) meta-topology but showed distinct molecular patterns. A high proliferation index (MIB-1 monoclonal antibodies) was associated predominantly with meta-topologies 1 (parieto-occipital neopallial) and 3 (temporal neopallial). Tumors mapping to meta-topology 3 (temporal neopallial) showed a preference for MGMT promoter methylation. IDH-1 mutations mapped strongly to meta-topology 4 (unisegmental) and, although less prominently, to meta-topology 2 (frontal neopallial) and 5 (allopallial). IDH-1 wildtype status was prominent in meta-topology 1 (parieto-occipital neopallial) and, to a lesser extent, in meta-topology 3 (temporal neopallial). 1p19q co-deletion status was associated with meta-topology 2 (frontal neopallial). WHO grade 3 gliomas mapped prominently to meta-topologies 2 and 4, i.e., the frontal neopallial and unisegmental types. Their contribution to meta-topology 5 (allopallial) and meta-topology 1 (parieto-occipital neopallial) was intermediate. WHO grade 2 glioma showed a similar pattern of factor contribution with dominance in meta-topologies 2 (frontal neopallial), 4 (unisegmental), and 5 (allopallial) and intermediate weight in meta-topology 1 (parieto-occipital neopallial).

**Figure 3.**
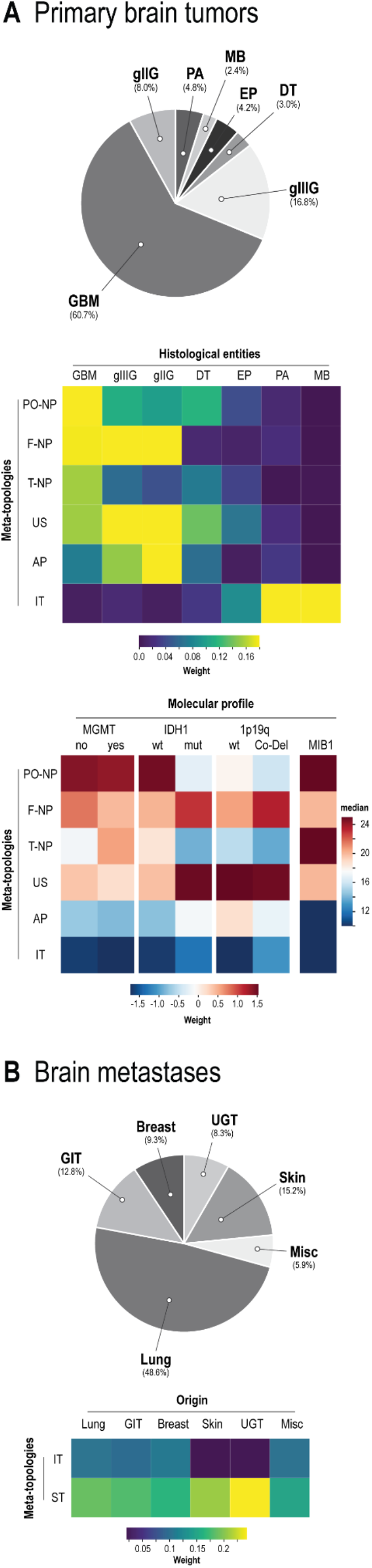
Histopathologic and molecular profiling of brain tumor meta-topologies. **A**. *Top:* Relative frequency of different histologic entities in the primary brain tumors cohort. *Upper matrix:* Mean expression of each meta-topology across primary brain tumor entities. *Lower matrix:* Summarized mean expression of each meta-topological molecular profiling based on mutational status (mean weight) and stratified median MIB-1/Ki-67 proliferation index. *Abbreviations:* 1p19q, 1p19q co-deletion; AP, allopallial; DT, developmental tumor; EP, ependymoma; gIIIG, WHO grade 3 glioma; F-NP, frontal neopallial; gIIG, WHO grade 2 glioma; GBM, glioblastoma; IDH1, Isocitrate dehydrogenase (IDH) mutation; IT, infratentorial; MB, medulloblastoma; MGMT, O(6)-methylguanine-DNA methyltransferase (MGMT) promoter methylation; MIB1, MIB-1/Ki-67 proliferation index; PA, pilocytic astrocytoma, PO-NP, parieto-occipital neopallial; T-NP, temporal neopallial; US, unisegmental. **B**. *Top:* Relative frequency of different brain metastases subtypes (depending on organ of origin). *Matrix:* Meta-topology specific enrichment (weight) of brain metastases subtypes. *Abbreviations (alphabetic order):* GIT, gastrointestinal tract (mouth, tonsil, parotid, esophagus, stomach, gallbladder, pancreas, colorectal cancer); IT, infratentorial, Misc, miscellaneous (cancer of unknown primary, adrenal, leukemia, sarcoma, mesothelial, thyroid); ST, supratentorial; UGT, urogenital tract (kidney, bladder; ovary, tube, uterus; testes, prostate).

Brain metastases most frequently arose from lung cancer (48.6%), melanoma (15.2%), or gastrointestinal cancer (12.8%) (Figure 3B). Brain metastases from lung, gastrointestinal or breast cancer contributed relevantly to both meta-topologies. However, there was a pronounced preference of brain metastases from melanoma and urogenital cancer to meta-topology 2, i.e., the supratentorial type.

### Brain tumor meta-topologies uncover survival differences

For survival analysis, patients were individually assigned to their highest expressed meta-topology (Supplemental Table 5-6). In primary brain tumors, meta-topology 6 (infratentorial) showed the highest survival probability (hazard ratio 0.12, 95% confidence interval: 0.07 to 0.23, p < 0.0001; Figure 4A, Supplemental Table 7A), thus supporting the NNMF results, which predominantly mapped ependymoma and pilocytic astrocytoma to this meta-topology. Poorest prognosis was seen in patients with tumors of meta-topologies 1 (parieto-occipital neopallial, reference for Cox proportional hazards analysis) or 3 (temporal neopallial, hazard ratio 1.07, 95% confidence interval: 0.77 to 1.49, p = 0.69) consistent with the histopathologic findings as both meta-topologies specifically mapped to glioblastoma. The analysis emphasizes the histopathologic- and anatomical-clinical link uncovered by the unsupervised and data-led analysis strategy. To capture hidden information beyond the histopathologic characteristics that might be directly linked to the clinical course, we adjusted for the histopathologic entities and showed that patients with tumors classified as meta-topology 4 (unisegmental) demonstrated a survival advantage (hazard ratio 0.65, 95% confidence interval: 0.49 to 0.87, p = 0.004). The survival analysis in glioblastoma (Figure 4B, Supplemental Table 7B) provided further evidence that patient with tumors matching to meta-topology 4 (unisegmental) show a better overall survival (hazard ratio 0.65, 95% confidence interval: 0.48 to 0.89, p = 0.007), underlining the anatomical-clinical link within histopathologically identical tumors. In contrast, brain metastases showed no evidence for a difference in survival between patients with assigned to the infra- or supratentorial meta-topology across all analyses (brain metastases overall unadjusted or adjusted for origin, and lung cancer brain metastases) (Figure 4C-D).

**Figure 4.**
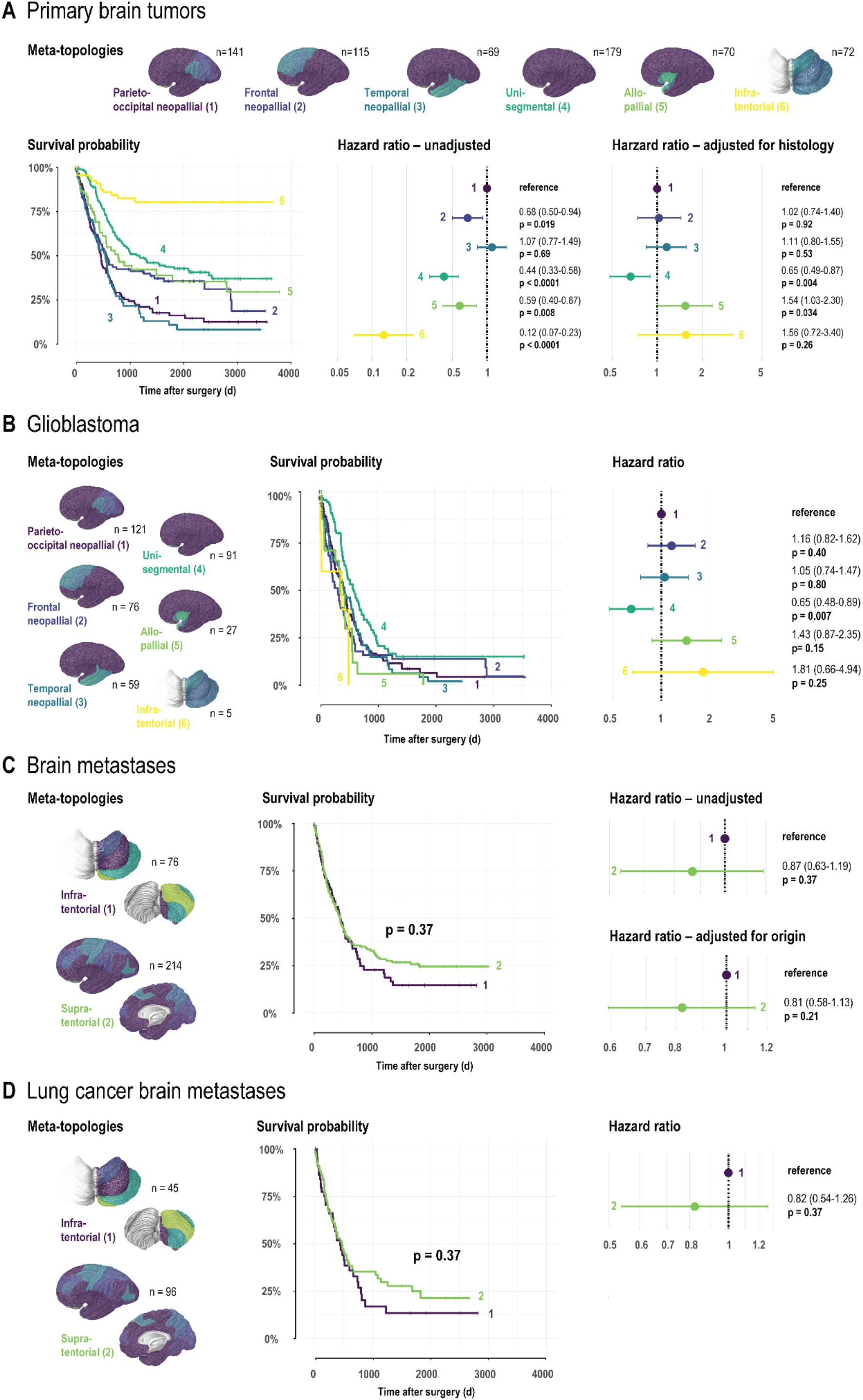
Brain tumor meta-topologies and patient survival. **A**. Survival in patients with primary brain tumors stratified by the corresponding meta-topology of highest relevance. *Left:* Kaplan-Meier curves. Risk tables and censoring events are given in Supplemental Figure 1A. *Right:* Forest plots providing meta-topology-specific hazard ratios with 95% confidence interval based on unadjusted and histology-adjusted Cox proportional hazards analyses. *Abbreviations:* d, days. **B**. Survival in patients with glioblastoma stratified by the corresponding meta-topology of highest relevance. *Left:* Kaplan-Meier curves. Risk tables and censoring events are given in Supplemental Figure 1A. *Right:* Forest plots providing meta-topology-specific hazard ratios with 95% confidence interval based on Cox proportional hazards analysis. **C**. Survival in patients with brain metastases stratified by the corresponding meta-topology of highest relevance. *Left:* Kaplan-Meier curves. Risk tables and censoring events are given in Supplemental Figure 1B. *Right:* Forest plots providing meta-topology-specific hazard ratios with 95% confidence interval based on unadjusted and origin-adjusted Cox proportional hazards analyses. **D**. Survival in patients with lung cancer brain metastases stratified by the corresponding meta-topology of highest relevance. *Left:* Kaplan-Meier curves. Risk tables and censoring events are given in Supplemental Figure 1B. *Right:* Forest plots providing meta-topology-specific hazard ratios with 95% confidence interval based on Cox proportional hazards analysis.

## Discussion

We present a novel data-led approach using machine learning to explore generalizable topological patterns across different entities of brain tumors. Using a quantitative out-of-sample evaluation strategy, we extracted six distinct meta-topologies in primary brain tumors and two meta-topologies in brain metastases in a cohort of 936 patients with fine-grained anatomical tumor annotations. The anatomical configuration of meta-topologies and their unique histopathologic profiles and prognoses provide insights into tumor biology and may enrich the current classification of brain tumors supporting more personalized clinical decision making.

The notion of latent meta-topologies with distinct histologic and molecular profiles in primary brain tumors emphasizes the concept of pathoclisis encountered in various other neurological disorders.^10–12^ The interpretation of their spatial architecture could enhance our biological understanding of tumor origin and evolution. First, all meta-topologies in primary brain tumors shared a transpallial character, i.e., all radial sectors between the cortex and ventricle were equally relevant. This was contrasted by the radial pattern in metastases with cerebellar (meta-topology 1) and cerebral (meta-topology 2) cortico-subcortical dominance and a ventriculopetal gradient. These findings are consistent with previous descriptions of the metastatic preference for the cortico-medullary boundary^29^ and a spatiotemporal behavior of primary brain tumors within ventriculocortical radial units.^8,9^ Second, except for meta-topology 4, the supratentorial meta-topologies identified in primary brain tumors demonstrated sharply defined gyral patterns, implying a segmental parenchymal growth behavior. Phylogenetic factors may contribute to segmental boundaries since meta-topology 5 mapped predominantly to allopallial structures (archipallium, paleopallium, and mesopallium), while meta-topologies 1-3 showed distinct neopallial patterns (parieto-occipital, frontal and temporal).^30^ In contrast, the parenchymal patterns in brain metastases corresponded to the border zones between the major cerebellar (meta-topology 1) and cerebral (meta-topology 2) arteries, consistent with previous descriptions of the metastatic tendency of origin in arterial watershed areas.^29^ Third, the meta-topologies in primary brain tumors were strongly defined by distinct ventricular segments, while the ventricles did not contribute relevantly to the meta-topologies in brain metastases. In contrast to earlier beliefs, niches of neuroepithelial stem cells seem to persist in the adult brain serving as a source of continuous cell replenishment.^31,32^ Such specialized niches have been identified in the dentate gyrus and the subventricular zone of the lateral ventricles.^33– 36^ It has been further hypothesized that glioblastoma, the most common primary brain tumor, may arise from such stem cell niches.^37^ This might also apply to other or even all tumors of neuroepithelial origin.^38,39^ Stem cell niches of distinct localizations could explain the association between the automatically extracted meta-topologies and specific segments of the ventricular system. The meta-topologies identified in this study were characterized in particular by the frontal horn, atrium, temporal horn/dentate gyrus, and the lateral recess of the 4th ventricle. These locations are reminiscent of the periventricular niches harboring stem cells and radial glia cells. Based on our patient-based findings, we thus hypothesize that specific radial ventriculo-cortical units are determined by their periventricular neuroepithelial stem cells and radial glial cells and potentially inform and shape the topological anatomy of primary brain tumors.^9,40,41^

Patients with primary brain tumors mapping to meta-topology 4 had a survival advantage after adjustment for tumor histology. This finding was confirmed within the group of patients with glioblastoma. Meta-topology 4 showed a transpallial character, comparable to the other supratentorial meta-topologies in primary brain tumors, but lacked a specific parenchymal pattern. The unigyral character of meta-topology 4 explains its lack of a specific gyral pattern, since topological analyses depend on the interrelationships between structures, but not their mere involvement.^13^ Meta-topology 4 rather constitutes a less advanced tumor stage than a separate entity with the potential to progress to a specific topological pattern. Meta-topology 4 was determined by a focal tumor contact to the ventricle wall, distinguishing it from the other supratentorial meta-topologies associated with diffuse ventricle wall involvement. The contact pattern to the ventricular system was shown to be a relevant prognostic factor and constitutes a cornerstone of a previously proposed anatomical staging of primary brain tumors.^9^

In brain metastases, melanoma and urogenital tract metastases contributed almost exclusively to the supratentorial meta-topology. The predominant supratentorial distribution is in accordance with previous studies suggesting a relative underrepresentation of melanoma metastases to the cerebellum.^42,43^ There was no evidence for a difference in survival between the infra- and supratentorial meta-topologies in brain metastases. The similarity may be explained by the fact that the presence of a specific brain tumor meta-topology is prognostically less critical than the stage and adjuvant treatment options of the underlying primary disease. The observation that meta-topologies do not offer a survival stratification in brain metastases is consistent with previous reports that the location of brain metastases is of minor overall prognostic significance.^44^

We introduce the concept of a data-driven analysis of topological classes in primary and secondary brain tumors. The proposed machine learning framework offers an unsupervised approach to identifying latent brain tumor meta-topologies in a data-centric fashion.^45^ That is, the algorithm learns without labels directly from the raw anatomical profiles. Notably, information about the individual histology, molecular pathology, or clinical parameters is at no stage available to the algorithm. Yet, we show that coherent and plausible patterns can be analytically retrieved based on the anatomical structure alone.

The addition of a macroscopic level to the current molecular and histological dimensions may complement primary brain tumor classification. The stratification by meta-topology – in addition to histological and molecular entities – offers new approaches to personalized medicine. Surgical or radiotherapeutic interventions may be tailored to the patient-specific expression of meta-topologies in primary brain tumors. Based on the hypothesis that primary brain tumors, given their neuroepithelial nature, orient along defined radial ventriculo-cortical units, local therapy could potentially target the entire affected anatomical segment. Meta-topologies may provide insights into the spatial behavior of primary brain tumors and argue for an segmentally tailored and functionally limited resection followed by focused irradiation^46^ rather than unselective supratotal resections, the latter being currently an emerging concept with limited evidence.^47–49^

## Data Availability

All data is available online at https://doi.org/10.5281/zenodo.5457402. Full codes are available online (https://doi.org/10.5281/zenodo.5515356).

https://doi.org/10.5281/zenodo.5515356

## Contributions

Conception and design of the study: KA, JMK, DD.

Data collection: KA, CS.

Analysis and/or interpretation of data: JMK, KA, DD.

Drafting the manuscript: KA, JMK, DD.

Review and revision of the manuscript: GN, HC, DB, SBE, VES, FV, MW, LR, CS, NK.

Approval of the version of the manuscript to be published: JMK, DD, GN, HC, DB, SBE, VES, FV, MW, LR, CS, NK, KA.

