## Supplemental for "Meta-topologies define distinct anatomical classes of brain tumors linked to histology and survival"

[1] Neurosurgical Artificial Intelligence Laboratory Aachen (NAILA), RWTH Aachen University Hospital, Aachen, Germany
[2] Department of Neurosurgery, Faculty of Medicine, RWTH Aachen University, Aachen, Germany
[3] Center for Integrated Oncology, Universities Aachen, Bonn, Cologne, Düsseldorf (CIO ABCD), Germany.
[4] Department of Biomedical Engineering, McConnell Brain Imaging Centre, Montreal Neurological Institute, Faculty of Medicine, School of Computer Science, McGill University, Montreal, Canada
[5] Mila - Quebec Artificial Intelligence Institute, Montreal, Canada
[6] Institute of Neuroscience and Medicine (INM-7), Research Centre Jülich, Jülich, Germany.
[7] Institute of Systems Neuroscience, Medical Faculty, Heinrich Heine University Düsseldorf, Düsseldorf, Germany.
[8] Department of Neurosurgery, Clinical Neuroscience Center, University Hospital and University of Zurich, Zurich, Switzerland
[9] Department of Neurology, Clinical Neuroscience Center, University Hospital and University of Zurich, Zurich, Switzerland
[10] Division of Pediatric Neurosurgery, University Children's Hospital, Zurich, Switzerland
* shared

*Running title:* Brain tumor meta-topologies

*Corresponding author:*
Kevin Akeret, MD
Department of Neurosurgery, Clinical Neuroscience Center
University Hospital and University of Zurich
Frauenklinikstrasse 10
CH-8091 Zurich, Switzerland

ORCID: 0000-0002-5946-4999

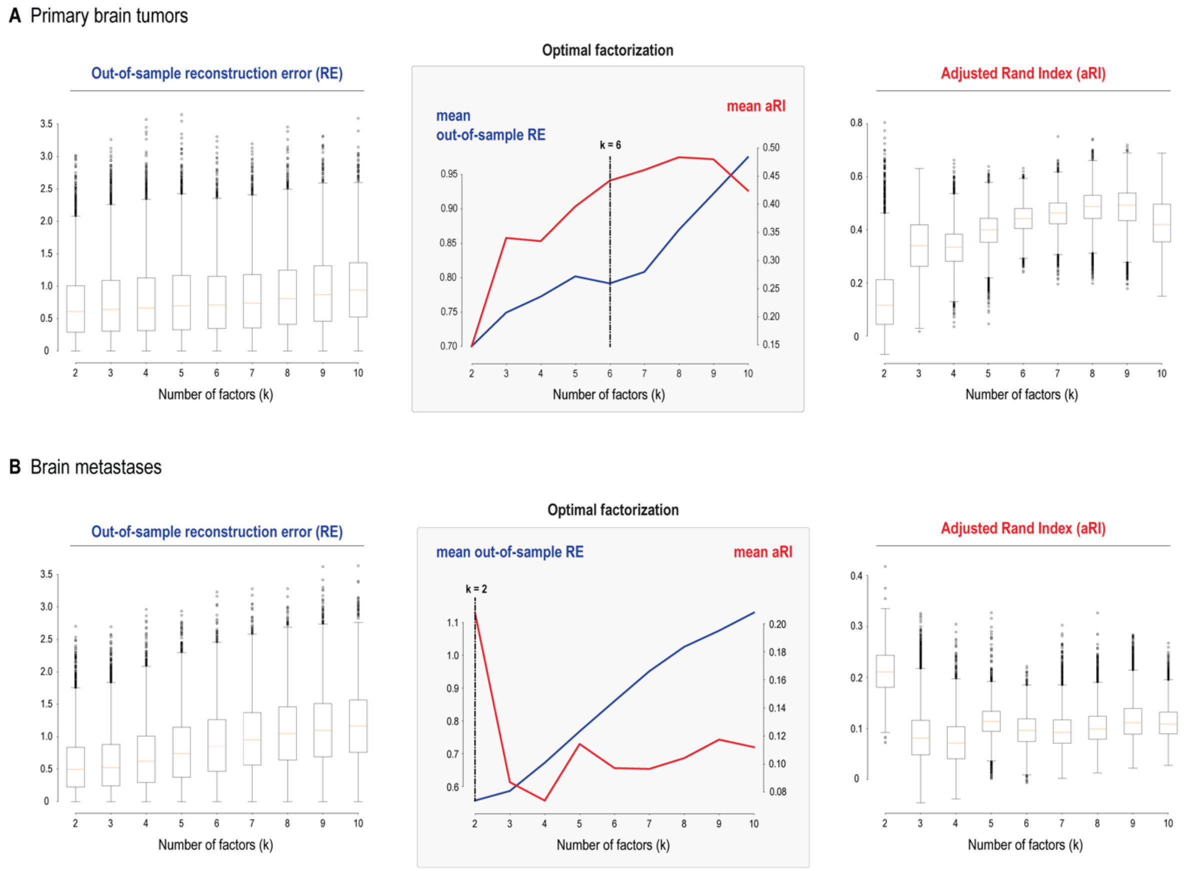


**Supplemental Figure 1. Optimal factorizations in primary and secondary brain tumors**

Quantitative evaluation of different non-negative matrix factorization (NNMF) solutions with *k* number of factors assessed in 10,000 bootstrapped split-half analyses by the out-of-sample increase in the reconstruction error (RE) and the adjusted Rand Index (aRI). Lower values for the out-of-sample increase in RE indicate better generalizability. Higher values of aRI indicate higher stability. **A** The optimal factorization in primary brain tumors resulted in *k*=6 meta-topologies. **B** The optimal factorization in brain metastases yielded *k*=2 meta-topologies.

A Primary brain tumors

B Brain metastases

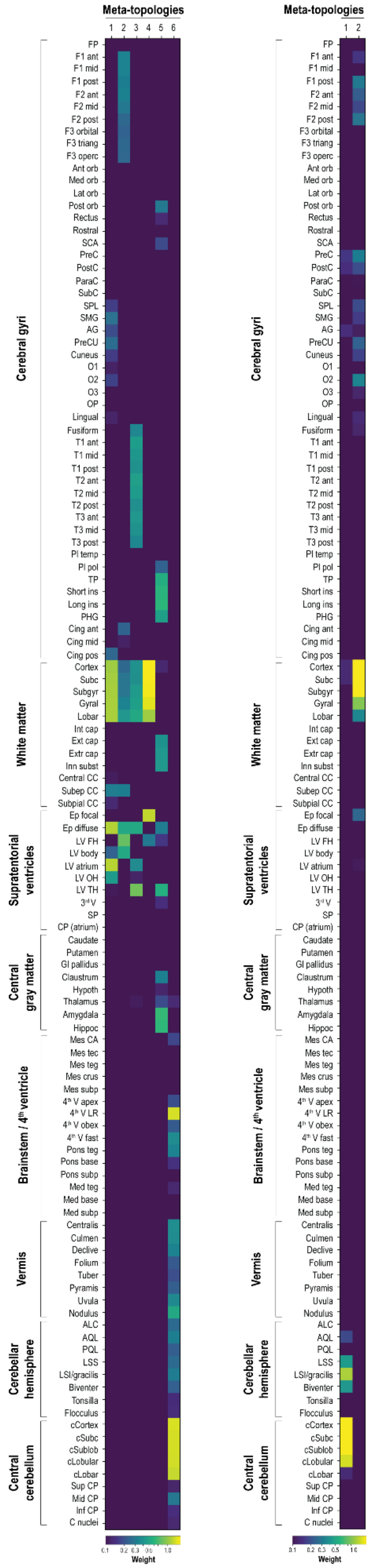

**Supplemental Figure 2. Detailed anatomy of primary and secondary brain tumor meta-topologies**

Supplement to Figure 1 and 2: Meta-topologies in primary brain tumors (**A**) and brain metastases (**B**) with their detailed underlying neuroanatomical distribution. *Abbreviations (in vertical descending order):* FP, frontal pole; F1, superior frontal gyrus; ant, anterior third; mid, middle third; post, posterior third; F2, middle frontal gyrus; F3 inferior frontal gyrus; Ant/Med/Lat/Post orb, anterior/medial/lateral/posterior orbital gyrus; SCA, subcallosal area; PreC, precentral gyrus; PostC, postcentral gyrus; ParaC, paracentral lobule; SubC, subcentral gyrus; SPL, superior parietal lobule; SMG, supramarginal gyrus; AG, angular gyrus; PreCU, precuneus; O1, superior occipital gyrus; O2, middle occipital gyrus; O3, inferior occipital gyrus; OP, occipital pole; T1, superior temporal gyrus; T2, middle temporal gyrus; T3, inferior temporal gyrus; Pl temp, planum temporale; Pl pol, planum polare; TP, temporal pole; ins, insular gyri; PHG, parahippocampal gyrus; Cingulate, cingulate gyrus; SubC, subcortical white matter sector; Subgyr, subgyral white matter sector; Gyral, gyral white matter sector; Lobar, lobar white matter sector; Int cap, internal capsule; Ext cap, external capsule; Extr cap, extreme capsule; Inn subst, innominate substance; CC, corpus callosum; Subep, subependymal; Ep, ependyma; LV, lateral ventricle; FH, frontal horn; OH, occipital horn; $3^{rd}$ V, third ventricle; SP, septum pellucidum; CP, choroid plexus; Gl, globus; Hypoth, hypothalamus; Hippoc, hippocampus; Mes, mesencephalon; CA, cerebral aqueduct (of Sylvius); tec, tectum; teg, tegmentum; subp, subpial; $4^{th}$ V, fourth ventricle; LR, lateral recess; fast, fastigium; ALC, ala lobuli centralis; AQL, anterior quadrangular lobule; PQL, posterior quadrangular lobule; LSS, superior semilunar lobule; LSI, inferior semilunar lobule; cCortex, cerebellar cortex; sSubc, cerebellar subcortical white matter sector; cSublob, cerebellar sublobular white matter sector; cLobular, cerebellar lobular white matter sector; cLobar, cerebellar lobar white matter sector; CP, cerebellar peduncle; C nuclei, cerebellar nuclei.

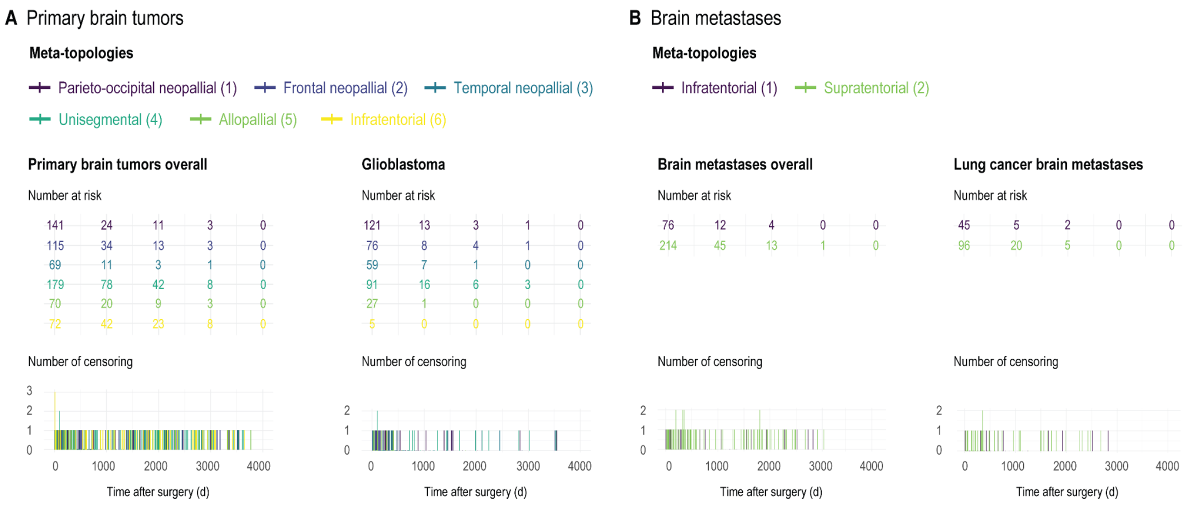


**Supplemental Figure 3. Risk tables and censoring events for the meta-topology-specific survival analyses**

Supplement to the Kaplan-Meier curves shown in Figure 4 providing the risk for an event over time (number at risk) and the number of censoring events over time (number of censoring) stratified by the corresponding meta-topology of highest relevance. **A.** Primary brain tumors overall (left) and glioblastoma (right). *Abbreviations:* d, days. **B.** Brain metastases overall (left) and lung cancer brain metastases (right).

### Supplemental Table 1. Cohort characteristics

The demographic, histopathologic, and clinical characteristics of the study cohort. *Abbreviations:* DT, developmental tumors; EP, ependymoma; gIIG, WHO grade II glioma; gIIIG, WHO grade III glioma; GBM, glioblastoma; GIT, gastrointestinal tract (mouth, tonsil, parotid, esophagus, stomach, gallbladder, pancreas, colorectal cancer); MB, medulloblastoma; Misc., miscellaneous (cancer of unknown primary, adrenal, leukemia, sarcoma, mesothelial, thyroid); PA, pilocytic astrocytoma; UGT, urogenital tract (kidney, bladder; ovary, tube, uterus; testes, prostate).
*based on immunohistochemistry or PCR

**Primary brain tumors**

| | Overall | GBM | gIIIG | gIIG | DT | EP | PA | MB |
|---|---|---|---|---|---|---|---|---|
| **n** | 646 | 379 | 105 | 50 | 19 | 26 | 30 | 15 |
| **Sex (male)** | 402 (62.2) | 244 (64.4) | 63 (60.0) | 31 (62.0) | 12 (63.2) | 14 (53.8) | 21 (70.0) | 9 (60.0) |
| **Age (years)** | 50.1 (21.6) | 60.8 (14.2) | 46.4 (15.7) | 43.1 (16.1) | 20.4 (15.7) | 24.6 (21.3) | 11.7 (10.4) | 16.6 (14.3) |
| **Karnofsky Performance Status** | 80.0 (12.6) | 76.9 (13.2) | 84.1 (10.0) | 86.7 (9.9) | 87.6 (8.3) | 86.7 (11.4) | 82.9 (9.2) | 83.0 (6.7) |
| **Modified Rankin Scale** | 1.7 (1.0) | 1.9 (1.0) | 1.4 (0.7) | 1.2 (0.7) | 1.1 (0.6) | 1.3 (0.9) | 1.5 (0.7) | 1.5 (0.5) |
| **Resection (vs. biopsy)** | 510 (78.9) | 285 (75.2) | 81 (77.1) | 37 (74.0) | 19 (100.0) | 24 (92.3) | 30 (100.0) | 14 (93.3) |
| **Chemotherapy** | 428 (69.1) | 269 (73.7) | 89 (86.4) | 35 (71.4) | 0 (0.0) | 13 (59.1) | 2 (7.1) | 14 (93.3) |
| **Radiotherapy** | 473 (76.2) | 311 (85.0) | 91 (88.3) | 29 (58.0) | 2 (10.5) | 17 (77.3) | 0 (0.0) | 15 (100.0) |
| **MIB1** | 25.0 (20.6) | 32.5 (19.2) | 18.7 (17.4) | 6.7 (4.8) | 3.4 (4.4) | 25.0 (24.4) | 3.8 (3.6) | 38.3 (26.5) |
| **1p19q co-deletion** | - | 2 (9.1) | 19 (23.8) | 21 (61.8) | - | - | - | - |
| **IDH1 mutation*** | - | 10 (3.4) | 45 (51.1) | 32 (72.7) | - | - | - | - |
| **MGMT promoter methylation** | - | 82 (38.0) | 9 (45.0) | 4 (57.1) | - | - | - | - |

**Metastases**

| | Overall | Lung | Skin | GIT | Breast | UGT | Misc. |
|---|---|---|---|---|---|---|---|
| **n** | 290 | 141 | 44 | 37 | 27 | 24 | 17 |
| **Gender (m)** | 149 (51.4) | 77 (54.6) | 24 (54.5) | 24 (64.9) | 1 (3.7) | 16 (66.7) | 7 (41.2) |
| **Age (years)** | 60.7 (11.9) | 61.4 (9.9) | 58.4 (13.6) | 65.6 (10.3) | 58.4 (12.5) | 56.2 (18.3) | 60.4 (10.3) |
| **Karnofsky Performance Status** | 76.9 (11.6) | 77.7 (11.0) | 79.8 (11.1) | 74.1 (10.4) | 77.8 (11.5) | 71.2 (15.1) | 75.9 (12.3) |
| **Modified Rankin Scale** | 2.0 (0.9) | 1.9 (0.8) | 1.8 (1.0) | 2.2 (0.8) | 1.9 (0.8) | 2.4 (1.0) | 1.9 (1.0) |
| **Resection (vs. biopsy)** | 277 (95.5) | 136 (96.5) | 43 (97.7) | 36 (97.3) | 27 (100.0) | 23 (95.8) | 12 (70.6) |
| **Chemotherapy** | 175 (62.3) | 94 (69.1) | 34 (77.3) | 12 (33.3) | 17 (65.4) | 10 (41.7) | 8 (53.3) |
| **Radiotherapy** | 255 (89.2) | 128 (92.1) | 42 (95.5) | 31 (83.8) | 25 (92.6) | 18 (75.0) | 11 (73.3) |

**Supplemental Table 2. Differential meta-topological anatomy in primary brain tumors**

Top ten anatomical items that define each meta-topology in primary brain tumors.

| Factor | Anatomical item | Weight | Factor | Anatomical item | Weight |
|---|---|---|---|---|---|
| **1** | Lateral ventricle – atrium | **1.19** | **4** | Cerebral cortex | **1.58** |
| | Ventricle wall – diffuse | **1.17** | | Cerebral subcortical white matter | **1.57** |
| | Cerebral gyral white matter | **1.11** | | Cerebral subgyral white matter | **1.54** |
| | Cerebral lobar white matter | **1.11** | | Cerebral gyral white matter | **1.41** |
| | Cerebral subgyral white matter | **1.10** | | Ventricle wall – focal | **1.25** |
| | Cerebral subcortical white matter | **1.09** | | Cerebral lobar white matter | **1.07** |
| | Cerebral cortex | **1.07** | | Lateral ventricle – frontal horn | **0.30** |
| | Lateral ventricle – occipital horn | **0.45** | | Subcentral gyrus | **0.09** |
| | Subependymal corpus callosum | **0.32** | | Short insular gyri | **0.06** |
| | Supramarginal gyrus | **0.28** | | Medial orbital gyrus | **0.06** |
| **2** | Lateral ventricle – frontal horn | **0.82** | **5** | Amygdala | **0.65** |
| | Lateral ventricle – body | **0.52** | | Hippocampus | **0.62** |
| | Ventricle wall – diffuse | **0.52** | | Long insular gyri | **0.61** |
| | Cerebral lobar white matter | **0.43** | | Short insular gyri | **0.60** |
| | Superior frontal gyrus – middle | **0.34** | | Lateral ventricle – temporal horn | **0.57** |
| | Superior frontal gyrus – anterior | **0.34** | | Temporal pole | **0.56** |
| | Cerebral gyral white matter | **0.34** | | Parahippocampal gyrus | **0.47** |
| | Middle frontal gyrus – anterior | **0.33** | | Innominate substance | **0.44** |
| | Subependymal corpus callosum | **0.32** | | Extreme capsule | **0.43** |
| | Superior frontal gyrus – posterior | **0.32** | | External capsule | **0.41** |
| **3** | Lateral ventricle – temporal horn | **0.86** | **6** | Cerebellar cortex | **1.37** |
| | Ventricle wall – diffuse | **0.53** | | 4th ventricle – lateral recess | **1.35** |
| | Cerebral lobar white matter | **0.52** | | Cerebellar sublobular white matter | **1.34** |
| | Middle temporal gyrus – anterior | **0.47** | | Cerebellar subcortical white matter | **1.34** |
| | Cerebral gyral white matter | **0.47** | | Cerebellar lobular white matter | **1.33** |
| | Superior temporal gyrus – anterior | **0.46** | | Cerebellar lobar white matter | **1.27** |
| | Middle temporal gyrus – middle | **0.45** | | Vermis – nodulus | **0.52** |
| | Cerebral subgyral white matter | **0.44** | | Vermis – centralis | **0.38** |
| | Inferior temporal gyrus – anterior | **0.42** | | 4th ventricle – fastigium | **0.38** |
| | Superior temporal gyrus – middle | **0.42** | | Vermis – culmen | **0.38** |

**Supplemental Table 3. Gyrality of primary brain tumors meta-topologies**

The gyral character of each meta-topology, i.e., the number (*n*) and proportion (*%*) of tumors without gyral involvement (none), those involving one gyrus (unigyral), and tumors involving multiple gyri (multigyral).

| | | Meta-topology | | | | | |
|---|---|---|---|---|---|---|---|
| | **Overall** | **1** | **2** | **3** | **4** | **5** | **6** |
| **n** | 646 | 141 | 115 | 69 | 179 | 70 | 72 |
| **Gyrality (%)** | | | | | | | |
| **none** | 119 (18.4) | 6 (4.3) | 17 (14.8) | 3 (4.3) | 0 (0.0) | 22 (31.4) | 71 (98.6) |
| **unigyral** | 285 (44.1) | 70 (49.6) | 32 (27.8) | 27 (39.1) | 147 (82.1) | 9 (12.9) | 0 (0.0) |
| **multigyral** | 242 (37.5) | 65 (46.1) | 66 (57.4) | 39 (56.5) | 32 (17.9) | 39 (55.7) | 1 (1.4) |

**Supplemental Table 4. Differential meta-topological anatomy in brain metastases**

Top ten anatomical items that define meta-topologies 1 and 2 in brain metastases.

| Factor | Anatomical item | Weight |
|---|---|---|
| **1** | Cerebellar cortex | **2.18** |
| | Cerebellar subcortical white matter | **2.17** |
| | Cerebellar sublobular white matter | **1.94** |
| | Cerebellar lobular white matter | **1.49** |
| | Inferior semilunar / gracile lobule | **1.24** |
| | Superior semilunar lobule | **0.47** |
| | Biventer lobule | **0.45** |
| | Anterior quadrangular lobule | **0.19** |
| | Precentral gyrus | **0.16** |
| | Postcentral gyrus | **0.15** |
| **2** | Cerebral subcortical white matter | **2.83** |
| | Cerebral cortex | **2.83** |
| | Cerebral subgyral white matter | **2.30** |
| | Cerebral gyral white matter | **1.03** |
| | Cerebral lobar white matter | **0.37** |
| | Middle occipital gyrus | **0.36** |
| | Precentral gyrus | **0.34** |
| | Superior frontal gyrus – posterior | **0.32** |
| | Middle frontal gyrus – posterior | **0.30** |
| | Ventricle wall – focal | **0.26** |

**Supplemental Table 5. Cohort characteristics in primary brain tumors stratified by dominant meta-topology**

The demographic, histopathologic, and clinical characteristics of the patients with primary brain tumors overall (A) and with glioblastoma (B) stratified by the dominant meta-topology. *based on immunohistochemistry or PCR

| Meta-topology | Overall | 1 | 2 | 3 | 4 | 5 | 6 |
|---|---|---|---|---|---|---|---|
| | | | Primary brain tumors overall (A) | | | | |
| n | 646 | 141 | 115 | 69 | 179 | 70 | 72 |
| Sex (male) | 402 (62.2) | 88 (62.4) | 64 (55.7) | 48 (69.6) | 119 (66.5) | 39 (55.7) | 44 (61.1) |
| Age (years) | 50.1 (21.6) | 59.6 (15.7) | 54.0 (18.1) | 57.7 (18.9) | 49.4 (18.5) | 50.4 (21.0) | 19.1 (18.6) |
| Karnofsky Performance Status | 80.0 (12.6) | 78.2 (11.5) | 75.8 (14.6) | 78.7 (12.5) | 84.8 (10.5) | 79.5 (12.1) | 80.4 (14.0) |
| Modified Rankin Scale | 1.7 (1.0) | 1.9 (0.9) | 2.0 (1.0) | 1.9 (1.0) | 1.3 (0.8) | 1.7 (0.9) | 1.7 (1.0) |
| Resection (vs. biopsy) | 510 (78.9) | 99 (70.2) | 75 (65.2) | 60 (87.0) | 171 (95.5) | 44 (62.9) | 61 (84.7) |
| Chemotherapy | 428 (69.1) | 100 (74.6) | 71 (64.5) | 50 (73.5) | 135 (78.0) | 40 (60.6) | 32 (47.1) |
| Radiotherapy | 473 (76.2) | 114 (84.4) | 81 (73.6) | 56 (82.4) | 144 (82.8) | 44 (66.7) | 34 (50.0) |
| MIB1 | 25.0 (20.6) | 28.0 (18.0) | 24.2 (18.2) | 29.6 (21.4) | 26.5 (22.1) | 16.3 (17.0) | 21.4 (24.3) |
| 1p19q co-deletion | 43 (31.4) | 3 (21.4) | 12 (52.2) | 0 (0.0) | 25 (34.7) | 3 (13.6) | 0 (0.0) |
| IDH1 mutation* | 88 (19.6) | 8 (7.5) | 17 (19.5) | 1 (2.0) | 53 (37.6) | 9 (20.5) | 0 (0.0) |
| MGMT promoter methylation | 95 (38.6) | 28 (37.3) | 18 (37.5) | 17 (47.2) | 24 (38.1) | 7 (36.8) | 1 (20.0) |
| | | | Glioblastoma (B) | | | | |
| n | 401 | 121 | 76 | 59 | 91 | 27 | 5 |
| Sex (male) | 244 (64.4) | 77 (63.6) | 45 (59.2) | 42 (71.2) | 62 (68.1) | 14 (51.9) | 4 (80.0) |
| Age (years) | 60.8 (14.2) | 62.7 (12.3) | 60.3 (14.7) | 62.4 (13.7) | 58.8 (15.4) | 62.2 (8.6) | 33.4 (25.1) |
| Karnofsky Performance Status | 76.9 (13.2) | 77.4 (12.0) | 72.0 (14.7) | 77.9 (12.8) | 81.2 (11.4) | 74.8 (12.2) | 60.0 (27.1) |
| Modified Rankin Scale | 1.9 (1.0) | 2.0 (1.0) | 2.2 (1.1) | 1.9 (1.0) | 1.6 (0.9) | 2.1 (0.9) | 3.2 (1.3) |
| Resection (vs. biopsy) | 285 (75.2) | 85 (70.2) | 47 (61.8) | 50 (84.7) | 87 (95.6) | 15 (55.6) | 1 (20.0) |
| Chemotherapy | 269 (73.7) | 86 (74.1) | 47 (64.4) | 45 (77.6) | 76 (85.4) | 13 (52.0) | 2 (50.0) |
| Radiotherapy | 311 (85.0) | 100 (85.5) | 56 (76.7) | 51 (87.9) | 81 (91.0) | 21 (84.0) | 2 (50.0) |
| MIB1 | 32.5 (19.2) | 30.6 (17.9) | 30.0 (17.2) | 32.8 (20.9) | 36.8 (20.6) | 31.7 (19.0) | 40.6 (26.7) |
| 1p19q co-deletion | 2 (9.1) | 0 (0.0) | 1 (50.0) | 0 (0.0) | 1 (14.3) | 0 (0.0) | 0 (-) |
| IDH1 mutation* | 10 (3.4) | 3 (3.2) | 4 (6.5) | 0 (0.0) | 3 (4.1) | 0 (0.0) | 0 (0.0) |
| MGMT promoter methylation | 82 (38.0) | 28 (38.4) | 14 (34.1) | 17 (48.6) | 17 (33.3) | 6 (42.9) | 0 (0.0) |

**Supplemental Table 6. Cohort characteristics in brain metastases stratified by dominant meta-topology**

The demographic, histopathologic, and clinical characteristics of the patients with brain metastases overall (A) and lung cancer brain metastases (B) stratified by the dominant meta-topology.

| | Metastases overall (A) | | | Lung cancer metastases (B) | | |
|---|---|---|---|---|---|---|
| **Meta-topology** | **Overall** | **1** | **2** | **Overall** | **1** | **2** |
| **n** | 290 | 76 | 214 | 141 | 45 | 96 |
| **Gender (m)** | 149 (51.4) | 41 (53.9) | 108 (50.5) | 77 (54.6) | 26 (57.8) | 51 (53.1) |
| **Age (years)** | 60.7 (11.9) | 58.5 (11.9) | 61.5 (11.8) | 61.4 (9.9) | 58.7 (9.7) | 62.6 (9.8) |
| **Karnofsky Performance Status** | 76.9 (11.6) | 77.5 (10.5) | 76.7 (12.0) | 77.7 (11.0) | 78.9 (10.3) | 77.2 (11.4) |
| **Modified Rankin Scale** | 2.0 (0.9) | 1.9 (0.8) | 2.0 (0.9) | 1.9 (0.8) | 1.8 (0.7) | 1.9 (0.9) |
| **Resection (vs. biopsy)** | 277 (95.5) | 72 (94.7) | 205 (95.8) | 136 (96.5) | 43 (95.6) | 93 (96.9) |
| **Chemotherapy** | 175 (62.3) | 49 (68.1) | 126 (60.3) | 94 (69.1) | 32 (72.7) | 62 (67.4) |
| **Radiotherapy** | 255 (89.2) | 67 (90.5) | 188 (88.7) | 128 (92.1) | 41 (93.2) | 87 (91.6) |

**Supplemental Table 7. Pairwise comparison of survival curves stratified by meta-topologies in primary brain tumor overall and glioblastoma**

Evidence for a difference between the meta-topology-stratified survival curves in (**A**) primary brain tumors overall and in (**B**) glioblastoma according to pairwise comparison with the log-rank test and Bonferoni-Holm correction for multiple testing. ****very strong evidence ($p < 0.001$),* ***strong evidence ($p < 0.01$),* **evidence ($p < 0.05$), +weak evidence ($p < 0.1$).*

| Meta-topology | 1 | 2 | 3 | 4 | 5 |
|---|---|---|---|---|---|
| **(A) Primary brain tumors overall** | | | | | |
| **2** | 0.041* | NA | NA | NA | NA |
| **3** | 0.68 | 0.031* | NA | NA | NA |
| **4** | <0.0001*** | 0.0085** | <0.0001*** | NA | NA |
| **5** | 0.013* | 0.51 | 0.0092** | 0.11 | NA |
| **6** | <0.0001*** | <0.0001*** | <0.0001*** | <0.0001*** | <0.0001*** |
| **(B) Glioblastoma** | | | | | |
| **2** | 0.58 | NA | NA | NA | NA |
| **3** | 0.81 | 0.76 | NA | NA | NA |
| **4** | 0.026* | 0.0076** | 0.03* | NA | NA |
| **5** | 0.4 | 0.76 | 0.4 | 0.0075** | NA |
| **6** | 0.43 | 0.75 | 0.4 | 0.074+ | 0.63 |
